# Gynaecological health patterns and motherhood experiences of female professional football players

**DOI:** 10.1101/2024.08.26.24312465

**Authors:** Dimakatso Ramagole, Dina C Janse van Rensburg, Charlotte Cowie, Ritan Mehta, Gopika Ramkilawon, Babette M. Pluim, Gino Kerkhoffs, Vincent Gouttebarge

## Abstract

**Aims:** To explore health patterns in female professional football players in the domains of gynaecological health patterns, contraceptive use, body perception and motherhood experiences, including return to play after childbirth.

**Methods:** An online questionnaire was emailed to active female professional football players via email. Participants were asked about their menstrual cycle,contraception use and motherhood. Validated questionnaires were used to assess body dissatisfaction (BD) and drive for thinness (DT).

**Results:** A total of 74 female professional football players were enrolled. The mean age at menarche was 13.5 years, average cycle length of 26 days and a bleeding period of 5 days. Cycle irregularities were experienced by 30% of participants, and menstrual symptoms by 74%. Half of the participants used contraceptives, with 60% using hormonal contraceptives, primarily oral contraceptive pills (38%), followed by implants (20%). Participants had a normal BD score, but a higher-than-expected DT score. The motherhood rate was low (1%), with normal conception, vaginal delivery, return to training after 6 weeks, and return to competition after 12 weeks.

**Conclusion:** Cycle irregularities are common in female professional football players, with a significant number of cycle-related symptoms. The majority on contraceptives preferred hormonal contraceptives, especially oral contraceptive pills (OCP) followed by implants, reflecting trends seen in elite athletes gynaecological health. While body satisfaction scores were normal, there was an unexpectedly high drive-for-thinness score, similar to that observed in lean or weight-category sports. The rate of motherhood was low, consistent with previous findings in professional football players.

**What is already known on this topic:**

- Elite female athletes have unique challenges related to their menstrual cycle and symptoms.
- Elite athletes use mostly hormonal contraceptives, predominantly the OCP.
- They often have to choose between professional sports and motherhood.

*What this study adds:* Our findings show some similarities to other to other elite female athletes: - 30% have menstrual irregularities.
- 74% have negative menstrual symptoms.
- Motherhood is low.
- Our participants have a high drive for thinness.

*How this study might affect research, practice or policy:*

- This descriptive study is a basis for more prospective and epidemiological research in this cohort that has been established.
- More research will be stimulated in the domains of menstrual symptoms and coping strategies, choice of contraception and motherhood.
- This study should also stimulate communications regarding player contracts and maternity benefits, and include after career support for female professional football players.

## INTRODUCTION

In recent years, there has been an increase in female participation in sports. Menstrual irregularities, delayed menarche, secondary amenorrhoea and disordered eating (DE) are common amongst female athletes and have been associated with disadvantages in the sporting career, as well as shortening the sporting career of endurance athletes.^1^ The menstrual cycle is an integral part of female athletes, and some earlier research suggested that early menarche (below age 12 years) has been associated with a higher risk of breast cancer due to the early onset of ovulatory cycles.^2^

Several surveys have investigated contraceptive use, including the methods chosen and the reasons behind their use.^3–4^ These surveys showed that more than 50% of their participants were not on contraceptives at all,^4^ and those who were on contraceptives predominantly used oral contraceptive pills (OCP).^3^

Motherhood experiences (ME) have unique challenges for female athletes, as highlighted in the article ‘We are not superhuman, we’re human.’^5^ Some of the concerns raised by athletes included: ‘training a new body’ postpartum; safe return to training; breastfeeding and exercise; and managing time between sports and motherhood. These athletes emphasised the need to develop best practices and secure funding for a safe return to play (RTP).^5–6^

Planned and well-structured physical activity under the guidance of an obstetrician-gynaecologist during and after pregnancy has been shown to benefit both physical and mental well-being. These benefits include a decrease in gestational diabetes, caesarean and operative vaginal deliveries, and a shorter post-partum recovery time.^7^

Several surveys highlighted the difficult choice between motherhood and elite sports participation, the need for careful planning of pregnancy and fertility,^8^ concerns about pregnancy disclosures and fear of discrimination, challenges in training the pregnant body safely, and the lack of a supportive network and funding during the pre- and post-natal periods.^5^

These interviews suggest that fewer than 50% of female professional football players have employment contracts comparable to those of their male counterparts. This lack of contractual security places female footballers at a disadvantage because they struggle to sustain themselves financially if they get injured or decide to start a family.^8^

Based on the considerations mentioned above, this study aimed to achieve two primary objectives: 1) To explore gynaecological health patterns in female professional football players, including the menstrual cycle, menstrual symptoms, contraceptive use and body perception; 2) To examine motherhood experiences, including methods of conception and delivery, and return-to-play/return to competition after childbirth.

## METHODS

### Study designs and ethical considerations

An observational prospective cohort study over a follow-up period of six months was conducted using the Strengthening the Reporting of Observational Studies in Epidemiology statement in order to guarantee the quality of reporting.^9^ The Medical Ethics Review Committee of the Amsterdam University Medical Centers (Amsterdam UMC, location AMC) provided ethical approval for the study (Drake Football Study: NL69852.018.19 | W19_171#B202169). The study was conducted in accordance with the Declaration of Helsinki (2013).

### Setting

Participants from English and French speaking countries were recruited by Football Players Worldwide (FIFPRO) and affiliated national unions via email from September 2020 to May 2021. Initial data collection was completed in December 2022, and participants will be followed at 6 months’ intervals over a period of 10 years.

### Participant selection

The study population consisted of female professional footballers. Inclusion criteria were: (a) being a professional footballer, (b) being female, (c) being able to read and comprehend texts in English or French. In our study, the definition for a professional footballer was that she (i) trains to improve performance, (ii) competes in the highest or second highest national league, and (iii) has football training and competition as a major activity (way of living) or focus of personal interest, devoting several hours in all or most of the days for these activities, and exceeding the time allocated to other types of professional or leisure activities. Sample size calculation indicated that at least 40 participants were needed to reach a power of 80% (confidence interval (CI) of 95% and absolute precision of 7%) under the assumption of an anticipated population proportion (prevalence) of 5%.^10^

### Dependent variables

Various female-specific variables were collected through single questions related to (history of) pregnancy, fertility (e.g., in vitro fertilisation, sperm donation), miscarriage, adoption, date of menarche, menstrual cycles, (history of) amenorrhoea, use of contraceptive measures (e.g., oral, implant), experiences related to motherhood (e.g., vaginal delivery, caesarean delivery), return to training (RTT)/return to competition (RTC) and body perception.

Body perception was measured by using the body dissatisfaction (BD) and drive for thinness (DT) scores. These were measured with two of the 12 primary scales of the Eating Disorder Inventory (EDI-3).^11^ This validated tool ensures a reliable interpretation of the psychometric test.^12^ BD relates to the disapproval of the overall shape of one’s body and the size of specific body regions of particular concern. DT consists of perceptual, behavioral, and attitudinal parts and is probably triggered when there is a discrepancy between actual and ideal body weight that exceeds the specific idealised preference of cultural thinness and involves body image dissatisfaction.^11^

The supplementary material presents all the questions in the survey. Annexure A.

### Descriptive variables

Participant characteristics and several descriptive variables (e.g., age, height, body mass index (BMI), player position, and stress fractures) were collected using an electronic questionnaire.

### Procedures and data storage

A baseline and a follow-up electronic questionnaire were set up in English and French (CastorEDC, CIWIT B.V, Amsterdam, the Netherlands), including all study outcome measures. In addition, the following descriptive variables were added to the baseline questionnaire: level of education, parallel activity (e.g., study, work), level of football, number of seasons as a professional footballer, history of hospitalisation, history of eating disorders, and smoking status. Information about the study was sent per email to potential participants by FIFPRO and affiliated national unions, and procedures were hidden from the principal researcher for privacy reasons. If interested in the study, all participants gave informed consent and received access to the baseline questionnaire. The follow-up questionnaire was sent six months later. Each questionnaire took about 15 minutes to complete. The responses to the questionnaires were coded and made anonymous for reasons of privacy and confidentiality. The electronic questionnaires were saved automatically on a secured electronic server that only the principal researcher could access. Players participated voluntarily in the study and were not rewarded for their participation.

### Statistical analyses

The Statistical software, R (version 4.2.1; http://www.r-project.org) was used to perform all data analyses. Descriptive analyses (mean, standard deviation (SD), frequency and proportions) were performed for all variables included in the study, including Body Mass Index (BMI) calculated as a ratio of weight/height squared.^13^

## RESULTS

### Participant characteristics

Seventy-four women athletes were recruited, with a mean age of 25 years and an average BMI of 22. Table 1 depicts demographics, football characteristics, career level and employment status. The majority of our participants were defenders (34%), followed by forwards (30%), midfielders (23%) and goalkeepers (14%). Most participants were did not have another paid job (70%), and most (87%) participated at the highest national level.

**Table 1:**
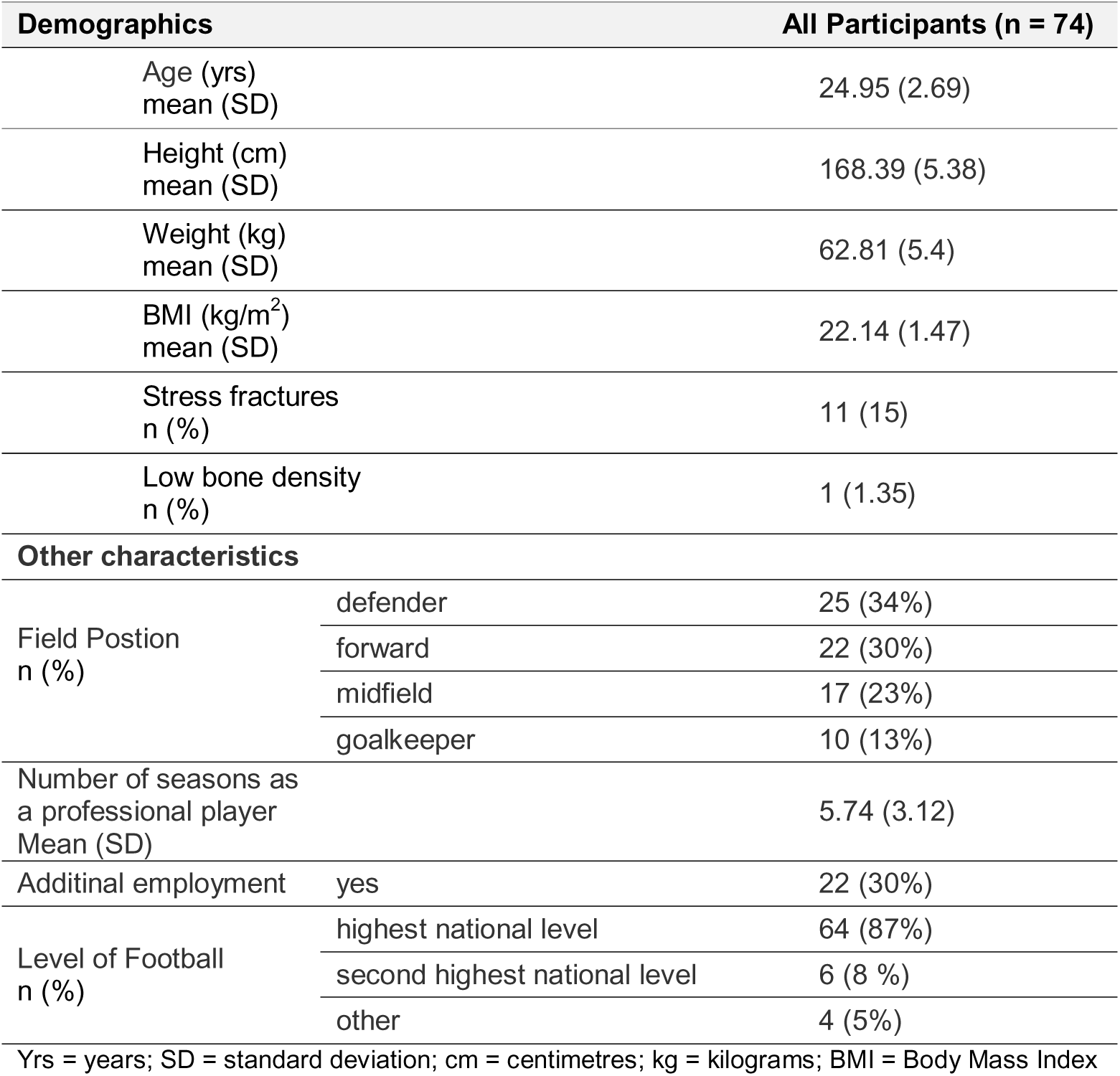
Demographics, football characteristics, employment status and career level.

### Gynaecological health patterns and body perception

Table 2 depicts gynaecological health patterns. The mean age at menarche was 13.5 years, with a menstrual cycle of 26 days and duration of menstrual bleeding of 5 days. Most participants reported pain during the bleeding period. Half of the participants used contraception.

**Table 2:** Gynaecological health patterns, BD and DT.

| Variable | Characteristics | All Participants (n = 74) |
| --- | --- | --- |
| Age of Menstruation (yrs)<br>Mean (SD) |  | 13.47 (1.3) |
| Duration of menstruation cycle (days)<br>Mean (SD) |  | 25.81 (13.09) |
| Duration of menstrual bleeding period (days)<br>Mean (SD) |  | 5.16 (3.42) |
| Description of menstruation cycle<br>n (%) | regular | 52 (70%) |
|  | irregular | 15 (20%) |
|  | very irregular | 7 (10%) |
| Pain during menstruation cycle<br>n (%) | during bleeding<br>period | 40 (54%) |
|  | none | 19 (26%) |
|  | before bleeding<br>period | 15 (20%) |
| Contraception Methods n (%) | none | 37 (50%) |
|  | pill | 14 (38%) |
|  | condom | 12 (32%) |
|  | other | 8 (22%) |
|  | implant | 7 (20%) |
|  | hormone<br>replacement | 4 (10%) |
|  | injection | 1 (2%) |
| Body dissatisfaction<br>Mean (SD) |  | 4.88 (5.08) |
| Drive for thinness<br>Mean (SD) |  | 3.59 (5.04) |

SD = standard deviation; BD = Body dissatisfaction; DT = Drive for thinness

### Motherhood experiences and RTT/RTC

Only one (1%) of the 74 participants had two children, both conceived naturally and delivered vaginally. Her RTT was 6 weeks, and RTC 12 weeks on both occasions.

## DISCUSSION

The findings from this study provide a baseline of active female professional football players’ gynaecological health behaviours, whilst most studies reported on retired elite athletes,^14^ or elite athletes from other sports.^15–16^ Our findings were that:1) hormonal contraceptives, particularly the oral contraceptive pill, were the most commonly used, 2) cycle irregularities were common, 3) most participants experienced cycle-related pain, and 4) motherhood experiences were rare during the active career.

### Menstrual cycle

Mernache is the average age at which menstrual bleeding starts in young females, and usually, this is between the ages of 12-13 years. It has been reported that the onset of menarche is usually delayed in athletes participating in lean sports, but no variations were noted between individual and team sports.^4^ Our participants had a mean menarche age of 13.5 years, comparable to other studies on elite athletes.^14^ Our results showed that 30% of our participants had cycle irregularities. Our findings indicate a higher prevalence of cycle irregularities compared to Swiss elite athletes from various sports, where approximately 15% reported such issues.^4^ However, our results are lower than those for elite athletes in Denmark^17^ and the United Kingdom,^15^ where 51% of athletes reported cycle irregularities. Retired semi- and professional football players reported a 40% prevalence of menstrual irregularities, with 22% experiencing amenorrhoea lasting longer than 3 months.^14^

Menstrual pain was reported by 74% of our participants. Symptoms were present before the bleeding period in 20% and during the menstrual phase in 54% of participants. This is comparable to the findings in a UK survey where 77% of elite athletes reported menstrual pain.^15^ Elite rugby players reported menstrual pain in 93% of participants,^16^ and 84% of elite Denmark athletes reported negative symptoms related to their menstrual cycle.^17^

A study among elite rugby and football players on premenstrual pain and coping strategies suggested a model including avoiding harm, awareness and acceptance, adjusting energy, self-care, and communication to help maintain well-being and physical activity during this period.^18^ Our study was not designed to investigate coping strategies, and this will require further investigation.

Several studies have shown that athletes believe that their performance declines when they experience menstrual-related symptoms.^16–17,19–20^ A survey of sub-elite female football players showed a reduction in endurance performance during the menstrual phase,^21^ while elite Australian football players reported higher perceived fatigue during this period.^22^ Our study did not investigate the impact of the menstrual cycle on exercise performance, highlighting the need for future research to address this gap in the literature on female professional football players.

### Contraceptive use

Our findings confirm that 50% of our participants were on contraceptives, mainly HCs (60%), predominantly the OCP (38%), followed by implants (20%) and injectable contraception (2%). These findings are consistent with findings amongst other elite athletes, with 50% on HCs, predominantly the OCP (68.%), followed by the implants (13%), injection (4%) and intra-uterine devices (3%).^15^ In elite Australian football codes, HCs use was 33%, with 81% on the OCP.^3^ A survey on Swiss players reported that 46% were on contraception, and 26% used the OCP,^4^ and a study on Danish players reported 57% use of HCs and all of them on OCP.^17^ Amongst retired professional football players from the United States (US), the trend was similar, although more participants (64.5%) had used HCs, predominantly the OCP (94.5%).^14^ Some of our participants reported the use of more than one contraceptive method. Our study was not designed to investigate reasons for the use of different methods and what ‘other methods’ were used. Further research is warranted on this topic.

### BD and DT

Our participants had a normal BD score of 5. This finding aligns with findings in federated Spanish amateur football players, who also reported high positive body images and a normal BD score of 5.^23^ We are not aware of other similar studies in female professional football players. A systematic review and meta analysis across different sports and genders highlighted the importance of sports participation in fostering positive body image. The study found that female athletes had higher BD scores than males, normal-weight athletes had higher BD scores than underweight ones, and compared to non-athletes, female athletes tend to have a higher body image perception.^24^

Our participants had a high mean DT of 4, which was not expected in football. Higher DT scores are common in aesthetic and weight category sports.^25^ We are not aware of other comparable studies in female professional football players. High DT has been correlated to susceptibility to musculoskeletal injuries due to low energy availability in the National Collegiate Athletic Association (NCAA) Division II female athletes and higher time loss from training.^26^ In this NCAA survey, 82% of participants sustained musculoskeletal injuries in-season. The mean number of injuries/training days loss was higher in the high DT group (over 2.0 ± 0.3), with approximately 9.8 days loss from training, whilst the low DT group (under 2.0 ± 0.3) had approximately 6.9 days loss.^26^

We found that 70% of our participants did not have another paid job, 58% were not studying, and 13 (17.6%) smoked. These factors, combined with a high DT, warrant further monitoring of these athletes to promote well-being. As confirmed in one study, high DT is a risk factor for the development of ED, and personal risk factors for DT include perfectionism, low self-esteem in sport, unemployment, and not studying^27^ and has been linked to tendencies towards smoking.^28^ Although ED was found to be higher in non-athletes compared to elite female football players,^29^ the combination of risk factors for ED and susceptibility to musculoskeletal injuries warrant further monitoring and research.

### Motherhood experiences

In our study, only one participant (1%) reported having had two pregnancies and she conceived naturally. Both children were delivered vaginally, and she returned to training and competition 6 and 12 weeks post-partum, respectively. The US study reported that 50% of the participants reported trying to fall pregnant, with a 94% success rate, mostly by intra-uterine insemination (51%), medication (49%) and in-vitro fertilisation (43%).^14^ Our sample is too small to compare. Still, we could assume that pregnancy during a professional football career remains challenging as players might be afraid to potentially jeopardise their career due to motherhood.

It has been shown that some professional athletes opt to choose between being mothers and continuing to participate in sports,^8^ but similar to our study, other athletes have successfully fallen pregnant and managed to RTT/RTC earlier than non-athletes.^30^ Although early RTT has been associated with an increased risk for stress fractures, high BD and high DT,^30^ this has not been confirmed in our study.

FIFPRO reported in 2017 that only 2% of female professional football players were mothers, and almost 45% expressed that they would have to leave the game early to start a family.^31^ Our study’s low percentage of motherhood is comparable to the FIFPRO findings. The choice between motherhood and professional football was not canvassed in our study, but it will form a basis for future research. FIFPRO has recently launched ‘The postpartum return to play guide’^32^ which details a 5-step guide beginning at the prenatal stage to return to exercise at 12 weeks postpartum, followed by football specific training, and finally high performance training.

### Strengths and Limitations

The strength of our study lies in its pioneering focus on the gynaecological health patterns and motherhood experiences of active female professional football players. Although our study is descriptive, our research provides a valuable cohort for future prospective and epidemiological studies, and implementation of the return to play guidelines after childbirth. We hope this study will inspire further research in the realm of female professional football players.

Our study has several limitations, including the small sample size and the lack of investigations into participants’ perceptions of exercise performance during the menstrual cycle, their coping strategies for menstrual symptoms, reasons for contraceptive choices, and their views on balancing motherhood with professional sport. We did not investigate any correlation between injuries and the menstrual cycle. Our findings on motherhood experiences are also limited by the low number of participants. Further research is needed to address these gaps.

### Implications

Women’s football is growing globally, and factors unique to women may affect their performance and/or shorten their sporting career. This study highlights the challenges faced by these athletes, including menstrual cycle-related symptoms that may affect performance and sporting career, and the importance of specialised gynaecological care for female elite athletes. Healthcare practitioners, coaches and team managers working with elite female athletes should be trained to improve communication, guide athletes on general trends in gynaecological health patterns, optimise well-being, and advise on safe RTT and RTC after motherhood.

More research, such as athlete perception of menstrual cycle effects on performance, coping strategies when experiencing menstrual pain, choice and reasons for use of contraceptives, and choice of sports versus motherhood, is needed in female professional football players.

## CONCLUSION

Cycle irregularities are common in female professional football players, with a significant number of cycle-related symptoms. The majority of those using contraceptives preferred hormonal contraceptives, predominantly oral contraceptive pills followed by implants, reflecting trends similar to other female elite athletes. While body satisfaction scores were normal, there was an unexpectantly high DT score, similar to that observed in lean or weight-category sports. The rate of motherhood was low, consistent with previous findings in professional football players.

## Data availability statement

All data relevant to the study are included in the article.

## PATIENT CONSENT FOR PUBLICATION

Not required.

## ACKNOWLEDGEMENTS

The authors would like to thank Football Players Worldwide (FIFPRO) and affiliated national unions for the support in recruiting professional footballers. The authors are grateful to all participants involved in the study.

## CONTRIBUTORS

All authors were involved in the design of the study. VG and GK were responsible for data collection. All authors were involved in the data analysis and data interpretation. DR drafted the manuscript, with critical review provided by all authors. All authors approved the final version of the manuscript.

## PATIENT AND PUBLIC PARTNERSHIPS

There are no patient and public partnerships to declare.

## ETHICAL APPROVAL

The Medical Ethics Review Committee of the Amsterdam University Medical Centers (Amsterdam UMC, location AMC) provided ethical approval for the study (Drake Football Study: NL69852.018.19 | W19_171#B202169). The study was conducted in accordance with the Declaration of Helsinki (2013).

